# Architectural Safety Mechanisms for Multi-Agent Clinical LLM Systems Under Knowledge Base Distribution Shift

**DOI:** 10.64898/2026.07.31.26359439

**Authors:** Muhammad Abiodun Sulaiman, Bolaji Fatai Oyeyemi, Hammed Sarafadeen

**Affiliations:** Department of Statistics, University of Ilorin, Ilorin, Nigeria; Department of Zoology, University of Ilorin, Ilorin, Nigeria; Department of Geography, University of Ilorin, Ilorin, Nigeria

**Keywords:** clinical decision support, dataset shift, multi-agent systems, retrieval-augmented generation, patient safety, LLM robustness

## Abstract

**Objective:** To evaluate whether multi-agent LLM architectures with explicit safety verification maintain guideline compliance when their clinical knowledge bases undergo temporal or institutional distribution shift.

**Materials and Methods:** We designed a controlled evaluation framework using 50,000 synthetic type 2 diabetes patients with CKD and hypertension comorbidities (500 per experimental condition). Four architecture modes (single-agent, naive RAG, linear multi-agent, stateful graph with safety floor) were tested under four shift regimes: baseline, temporal drift (updated eGFR thresholds), institutional vocabulary transformation (11 term-pair substitutions producing 0.36 cosine similarity degradation), and metadata erasure. The clinical task was medication reconciliation with contraindication detection. Two embedding models (all-MiniLM-L6-v2, PubMedBERT) and two LLM backends (Llama3-8B, Mistral-7B) were compared.

**Results:** Under institutional vocabulary shift, the linear pipeline’s Guideline Compliance Score dropped from 1.00 to 0.36 because retrieval degradation rendered critical contraindication guidelines unretrievable. The stateful graph architecture maintained GCS = 1.00 across all shift conditions through its regime-aware safety floor, which operates independently of retrieval quality. This pattern held across both LLM backends and both embedding models. The safety mechanism added 32.2s latency per patient under shift versus 12.5s for single-agent mode.

**Discussion:** Architectural choice (specifically whether audit findings are routed back to the summary agent) determines compliance under shift more than retrieval quality or model scale. The safety floor’s value is compliance maintenance, not semantic fidelity improvement.

**Conclusion:** Stateful multi-agent graphs with programmatic safety floors bound error propagation under clinical knowledge shift. The framework is reproducible on consumer hardware with no external API dependencies.

**Lay Summary:** When AI systems help doctors review medications, they rely on up-to-date medical guidelines stored in a database. If those guidelines change (because recommendations are updated or a hospital uses different terminology) the AI can silently give outdated advice. We tested whether connecting multiple AI agents in a loop, where one agent checks another’s work against safety rules, prevents this problem. It does: even when the database becomes unreliable, the safety-checking agent catches dangerous advice before it reaches the doctor. The trade-off is that the system takes about 20 extra seconds per patient.

## 1 Introduction

Deployed clinical AI systems increasingly operate as multi-step pipelines: parsing electronic health records, retrieving institutional guidelines, and generating treatment recommendations [1, 2, 3]. Singhal et al. showed that large language models can encode clinical knowledge at expert level on medical benchmarks, but Wornow et al. [4] argue that these results do not transfer reliably to production EHR settings where the data differs from training distributions. Unlike static classification models, agentic workflows depend on external knowledge bases that evolve independently of the model’s parameters [5, 6].

The challenge is that clinical knowledge bases shift. Prescribing guidelines update (temporal shift), institutions adopt different nomenclature for the same drugs and tests (institutional shift), and documentation structure changes as EHR systems are migrated (schema shift) [7, 8, 9]. When retrieval-augmented generation (RAG) systems encounter shifted knowledge, they can produce outputs that sound clinically plausible but reference outdated or locally incorrect guidance [10, 11, 12].

Multi-agent architectures have been proposed as a mitigation: specialized sub-agents (parsing, retrieval, auditing) connected by directed graphs, with verification loops that catch errors before they reach clinical users [13, 14, 15]. Constitutional AI [16] demonstrated that self-critique loops reduce harmful outputs without human labels, using principle-based evaluation. Our safety floor follows a similar logic, applied to factual correctness rather than harmfulness: a programmatic auditor evaluates clinical outputs against deterministic constraints, routing non-compliant outputs back for revision. However, no empirical evaluation has tested whether these mechanisms maintain compliance under controlled knowledge-base perturbations, or whether they merely add latency without safety benefit [17, 18].

### Objective

We evaluate whether a stateful multi-agent graph with a programmatic safety floor maintains guideline compliance under temporal, institutional, and schema shift conditions, compared to single-agent, naive RAG, and linear multi-agent baselines.

## 2 Background

### Medication errors in CKD populations

Patients with chronic kidney disease on metformin face a specific, well-documented risk: lactic acidosis when renal clearance drops below prescribing thresholds [19]. In a community-based cohort of 75,413 diabetic patients, Lazarus et al. [20] found that metformin was not associated with acidosis at eGFR 30–44 mL/min (adjusted HR 1.09, 95% CI 0.83–1.44), but carried a twofold risk increase below eGFR 30 (HR 2.07, 95% CI 1.33–3.22). The clinical significance of this threshold is precisely what makes guideline updates dangerous when knowledge bases go stale: a system trained on the older 30 mL/min threshold will not flag patients in the 30–45 range when the guideline shifts upward.

Afkarian et al. [21] found that 23% of T2DM patients with eGFR below 30 remained on metformin at their last clinic visit, pointing to persistent prescribing inertia despite clear contraindication. Pharmacist-led medication reconciliation reduces adverse drug event-related hospital revisits by 67% (pooled RR 0.33, 95% CI 0.20–0.53) across 17 studies involving 21,342 patients [22]. Cochrane reviews confirm that medication reconciliation reduces error rates, though effects on patient-level outcomes remain uncertain at larger scales [23]. For patients on dialysis specifically, medication reconciliation is considered foundational to safety given the complexity of their regimens [24].

### Dataset shift in clinical AI

Temporal shift degrades both discrimination and calibration in deployed clinical prediction models [7, 25]. Davis et al. [9] developed a system for detecting calibration drift, showing that clinical models deteriorate as environments change even when their discrimination remains stable. Subbaswamy and Saria [6] formalize causal approaches to shift-stable models. Our work extends this literature from predictive models to generative multi-agent systems, where shift manifests in the external knowledge base rather than in the input distribution. The distinction matters: in a predictive model, shift changes the relationship between features and outcome. In a generative RAG system, shift changes what the model retrieves, not what it computes.

### Multi-agent LLM systems

Recent surveys formalize multi-agent coordination patterns and highlight reliability challenges in LLM-based agent systems [14, 18, 15]. We contribute the first empirical stress-test of these architectures under controlled knowledge-base perturbations, as opposed to static benchmark evaluation.

### RAG in clinical settings

RAG has been applied to clinical decision support systems backed by institutional guideline databases [13, 1, 2]. Xiong et al. [26] benchmarked RAG across six LLMs for medical question answering, showing that retriever choice swings accuracy by up to 18 percentage points and that combining multiple medical corpora with multiple retrievers yields the best results. Thirunavukarasu et al. [11] review LLM applications in medicine broadly, noting that retrieval grounding reduces hallucination but does not eliminate it. What remains uncharacterized are the failure modes of clinical RAG under institutional vocabulary shift, where the same clinical concepts are expressed in incompatible terminology across hospitals. Our framework directly simulates this scenario.

## 3 Materials and Methods

### 3.1 Clinical task definition

The target clinical task is **medication reconciliation with contraindication detection**: given a patient’s current medications, diagnoses, and renal function (eGFR), generate a summary that identifies active contraindications and recommends appropriate action (discontinue, switch, or flag for pharmacist review). This task is directly relevant to medication safety in type 2 diabetes with chronic kidney disease, where metformin contraindication thresholds have changed over time [27]. Sutton et al. [28] reviewed clinical decision support systems, finding that benefits depend critically on knowledge currency and integration with clinical workflow; outdated rules produce alert fatigue and erode clinician trust.

### 3.2 Synthetic patient corpus

We constructed *ClinicalShift-2026* : 50,000 synthetic longitudinal patient profiles with type 2 diabetes mellitus (T2DM), stochastic comorbidity assignment (CKD stage 3: 32%, CKD stage 4: 20%, hypertension: 60%), and medication assignment based on clinical rules with a 5% prescribing-error rate simulating documented real-world inertia. Table 1 validates our cohort against published T2DM+CKD epidemiology. For each experimental condition, 500 patients were sampled via stratified random sampling (seed=42).

**Table 1:** Synthetic cohort characteristics compared to published T2DM+CKD epidemiology.

| Parameter | Our Cohort | Published | Source |
| --- | --- | --- | --- |
| Mean age (years) | 60 (SD 12) | 60–65 | UKPDS, ADVANCE |
| CKD stage 3 prevalence | 32% | 25–40% | (author?) [21] |
| CKD stage 4 prevalence | 20%<sup>†</sup> | 5–15% | (author?) [27] |
| HTN comorbidity | 60% | 70–80% | (author?) [27] |
| Metformin use | 85%<sup>†</sup> | 60–80% | Clinical practice |

<sup>†</sup>Deliberately enriched to ensure statistical power for safety-metric evaluation.

#### Why synthetic data

Controlled perturbation experiments require that the same patient population is exposed to different knowledge-base states, with all other variables held constant. This is not achievable with real clinical data: you cannot retroactively change a hospital’s guidelines and re-observe outcomes [29]. Synthetic generation gives us exact control over which guidelines are shifted and by how much, while keeping patient demographics, medications, and lab values fixed across conditions. The trade-off is realism. Synthetic notes lack the noise, abbreviations, and incompleteness of real EHR text. We accept this trade-off because our research question is about architecture, not about parsing quality, and we isolate parsing by using a deterministic rule-based extractor.

### 3.3 Multi-agent architecture

We model the clinical workflow as a stateful directed graph *G* = (*V, ε, S*) over an append-only global state:

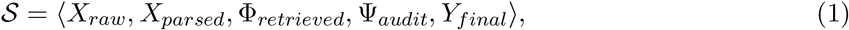

where *X*_*raw*_ is unstructured patient input, *X*_*parsed*_ is a structured clinical schema, Φ_*retrieved*_ are retrieved guidelines, Ψ_*audit*_ contains auditor findings, and *Y*_*final*_ is the output medication reconciliation summary. Each agent node *v*_*i*_ acts as a state transition operator: *v*_*i*_ : *S*_*t*_ *→ S*_*t*+1_.

Table 2 specifies what information each architecture mode provides to the summary agent and whether audit findings are actionable.

**Table 2:** Architecture mode specifications: information flow and safety mechanisms.

| Mode | Input to summary | RAG context | Audit feedback | Safety loop |
| --- | --- | --- | --- | --- |
| Single | $X_{raw}$ only | None | None | No |
| Naive RAG | $X_{raw} + \Phi_{baseline}$ | Baseline KB (stale) | Post-hoc only | No |
| Linear | $X_{raw} + \Phi_{shifted}$ | Shift-aware KB | Post-hoc only | No |
| Graph | $X_{raw} + \Phi_{shifted} + \Psi_{audit}$ | Shift-aware KB | Iterative | Yes ($L_{max}=4$) |

#### Sub-agent roles

The **Clinical Parser** (*v*_*parse*_) extracts diagnoses, medications, dosages, and eGFR from *X*_*raw*_ into *X*_*parsed*_. In this study, the parser is implemented as a deterministic rule-based extractor (no LLM involvement), isolating the effects of retrieval shift from parser errors. The **RAG Agent** (*v*_*rag*_) retrieves top-5 guidelines from a ChromaDB vector database via cosine similarity. The **Clinical Auditor** (*v*_*audit*_) evaluates *X*_*parsed*_ against regime-specific clinical rules (eGFR thresholds that vary by temporal epoch) to detect contraindications.

#### Auditor rule specification

The auditor applies deterministic threshold rules that are parameterized by the active regime:

- **Baseline/Institutional/Schema**: metformin contraindicated if eGFR *<* 30 mL/min
- **Temporal drift**: metformin contraindicated if eGFR *<* 45 mL/min (updated 2026 guideline)

The auditor operates on structured patient data (*X*_*parsed*_) against these deterministic rules, independent of retrieved guideline text. Conflicting retrieved documents do not affect the auditor’s verdict; it evaluates patient state against rules regardless of textual contradictions in Φ_*retrieved*_.

#### Safety floor mechanism

A compliance function *f*_*safe*_(Ψ_*audit*_) *→* {0, 1} gates the transition to output generation. If *f*_*safe*_ = 0 (contradictions detected), the state routes back for iterative correction (convergence bound *L*_*max*_ = 4). At *L*_*max*_, the system emits a “flagged-but-finalized” output that explicitly acknowledges unresolved contradictions rather than suppressing them silently.

### 3.4 Shift perturbation regimes

We apply four controlled perturbation regimes to the RAG agent’s knowledge base:

1. **Baseline** 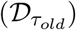: Unperturbed 2025 clinical guidelines (reference condition).
2. **Temporal drift** 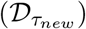: 18/60 documents (30%) modified with raised eGFR threshold (30*→*45 mL/min) and updated first-line therapy (SGLT2 inhibitors promoted).
3. **Institutional vocabulary swap** 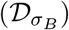 : 11 systematic term-pair transformations (e.g., “metformin”*→*”biguanide class agent”, “eGFR”*→*”estimated renal clearance rate”) preserving identical clinical content.
4. **Schema erasure**: All structured metadata (tags, epoch labels) stripped, forcing text-only retrieval.

#### Precondition validation

We validated that shift mechanisms produce measurable retrieval degradation before running the full experiment. Cosine similarity between query “metformin eGFR CKD contraindication” and the critical contraindication guideline: baseline = 0.63, institutional swap = 0.27 (drop of 0.36, below distractor-level relevance at 0.23).

### 3.5 Evaluation metrics

- **Semantic Fidelity Index (SFI):** BERTScore F1 [30] between output and ground-truth summary, using `deberta-xlarge-mnli`. 95% bootstrap CIs (1,000 resamples).
- **Fact-Checking Coverage Ratio (FCCR):** Per-patient fraction of ground-truth contradictions detected: 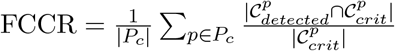
- **Guideline Compliance Score (GCS):** Binary: does the output acknowledge regime-dependent contraindications? Computed via deterministic string matching (“avoid”, “contraindicated”, “discontinue”, “switch” near drug name).
- **Temporal Calibration Agreement (TCA):** Alignment between verbalized confidence and epoch-correctness: 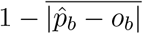 across confidence deciles [31].
- **Operational latency:** End-to-end wall-clock time per patient.

### 3.6 Experimental design

#### Embedding model comparison

To address whether our findings depend on the choice of embedding model, we compare `all-MiniLM-L6-v2` (general-purpose, 384d) against `PubMedBERT` (biomedical domain-specific, 768d) under institutional shift. We hypothesize that PubMedBERT’s vocabulary robustness reduces retrieval degradation but does not eliminate the need for the safety floor.

#### LLM backend comparison

To assess generalizability across model architectures, we compare Llama3-8B (4.7 GB, Q4 K M) and Mistral-7B-Instruct (4.1 GB, Q4 K M) under institutional shift for linear and graph modes.

### 3.7 Computational environment

All experiments were executed on a single Apple M3 Pro workstation (36 GB unified memory, 12 CPU, 18 GPU cores) running Ollama 0.30.10. Three concurrent workers (empirically optimal for this hardware). BERTScore computed on MPS backend. Zero external API calls. The full experiment is reproducible via `make all` on comparable hardware.^1^

## 4 Results

Table 3 summarizes performance across four architecture modes under three shift regimes using the default configuration (Llama3-8B, all-MiniLM-L6-v2).

**Table 3:** Main results: performance under shift (Llama3-8B, MiniLM embeddings, 500 patients/condition, 95% bootstrap CIs).

| Mode | Regime | SFI [CI] | GCS [CI] | TCA | Lat.(s) |
| --- | --- | --- | --- | --- | --- |
| Single | Baseline | 0.785 [0.781, 0.788] | 0.00 | 0.72 | 14.4 |
| Single | Institutional | 0.784 [0.780, 0.788] | 0.00 | 0.71 | 12.5 |
| Single | Temporal | 0.783 [0.779, 0.786] | 0.00 | 0.75 | 12.7 |
| Naive RAG | Temporal | 0.637 [0.634, 0.641] | 0.92 [0.88, 0.96] | 0.82 | 34.5 |
| Linear | Baseline | 0.639 [0.636, 0.642] | 1.00 | 0.86 | 29.1 |
| Linear | Institutional | 0.626 [0.623, 0.629] | 0.36 [0.23, 0.48] | 0.57 | 31.9 |
| Linear | Temporal | 0.636 [0.633, 0.638] | 1.00 | 0.80 | 26.5 |
| Graph | Baseline | 0.639 [0.636, 0.642] | 1.00 | 0.76 | 27.3 |
| Graph | Institutional | 0.627 [0.624, 0.630] | 1.00 | 0.69 | 32.2 |
| Graph | Temporal | 0.639 [0.636, 0.642] | 1.00 | 0.77 | 25.9 |

Under institutional vocabulary shift, the linear pipeline’s GCS drops from 1.00 to 0.36 (95% CI [0.23, 0.48]), while the graph architecture maintains GCS = 1.00 across all conditions. Single-agent mode achieves zero compliance by construction (no auditor). Schema erasure matches baseline for all modes, confirming retrieval depends on text content, not metadata structure (omitted for space). Figure 1 visualizes this pattern across all four architecture modes.

**Figure 1:**
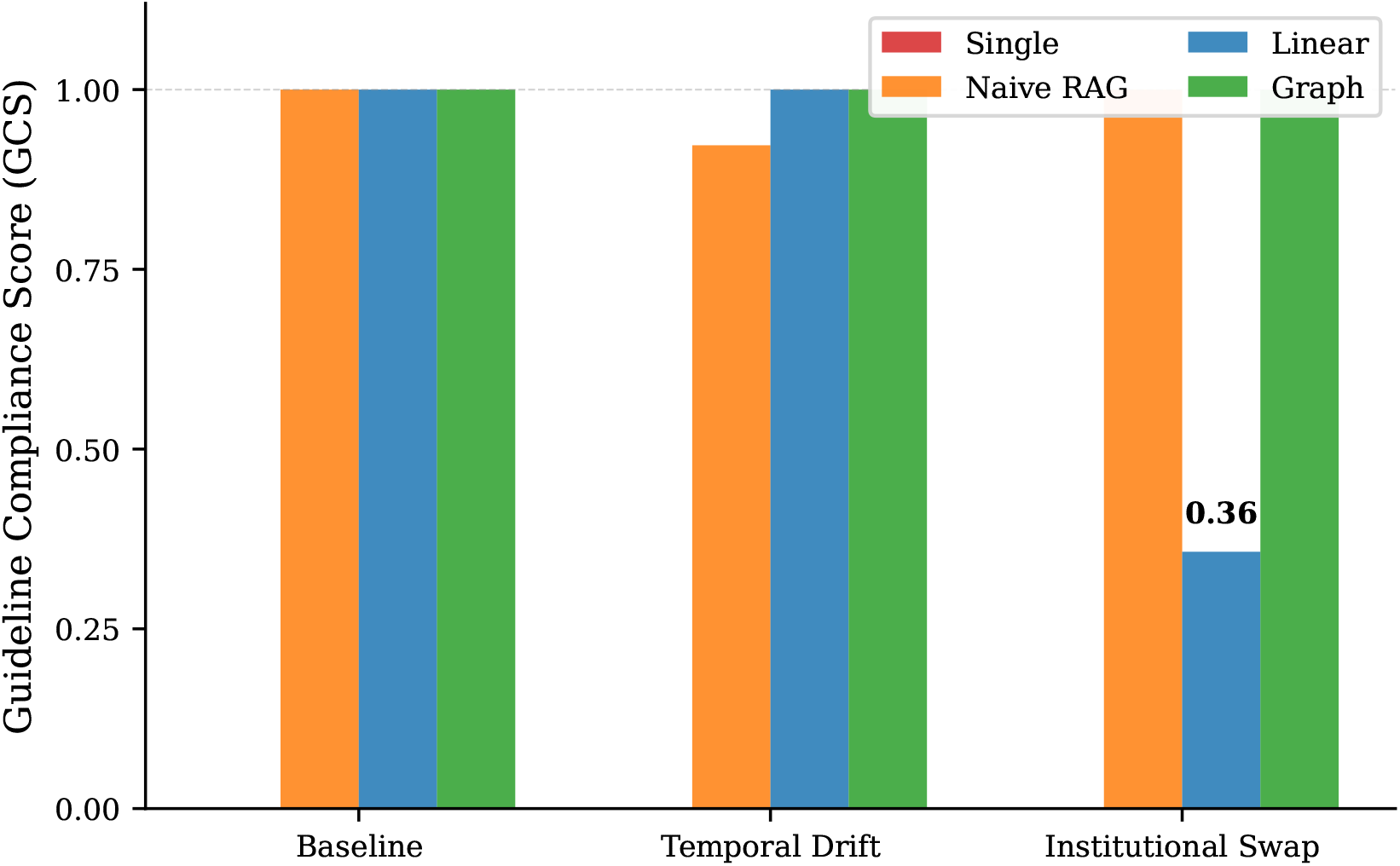
Guideline Compliance Score across architecture modes and shift regimes. Under institutional vocabulary shift, the linear pipeline drops to GCS = 0.36 while the graph architecture maintains 1.00. Single-agent mode has no auditor and scores zero by construction.

### 4.1 Embedding model comparison

Table 4 compares general-purpose (MiniLM) and biomedical (PubMedBERT) embeddings under institutional shift.

**Table 4:**
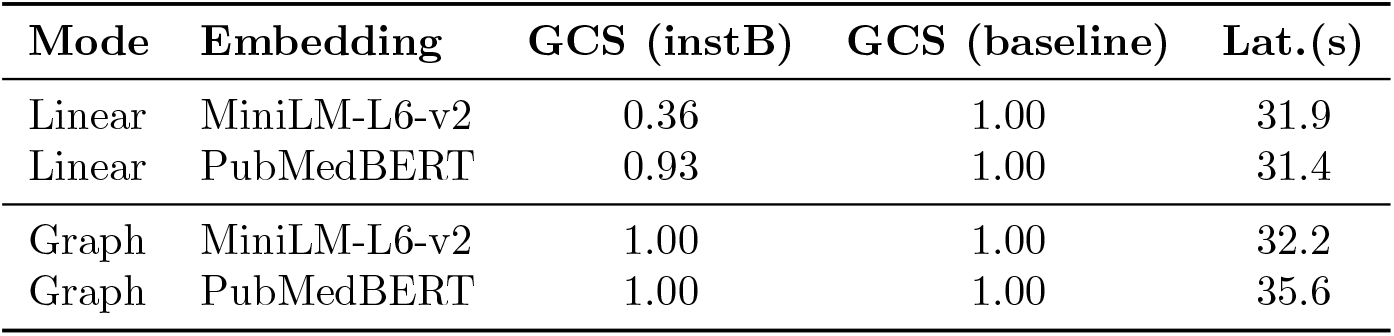
Effect of embedding model on compliance under institutional vocabulary shift.

PubMedBERT raises linear-mode GCS from 0.36 to 0.93 under institutional shift, demonstrating that domain-specific embeddings substantially reduce retrieval vulnerability. However, 7% of contraindicated patients are still missed. The biomedical model’s vocabulary robustness is not perfect. The graph architecture achieves GCS = 1.00 with both embedding models: the safety floor catches the residual failures that even PubMedBERT cannot prevent.

### 4.2 LLM backend comparison

Table 5 compares Llama3-8B and Mistral-7B under institutional shift.

**Table 5:** Effect of LLM backend on compliance under institutional vocabulary shift.

| Mode | LLM | GCS (instB) | GCS (baseline) | Lat.(s) |
| --- | --- | --- | --- | --- |
| Linear | Llama3-8B | 0.36 | 1.00 | 31.9 |
| Linear | Mistral-7B | 0.46 | 1.00 | 41.6 |
| Graph | Llama3-8B | 1.00 | 1.00 | 32.2 |
| Graph | Mistral-7B | 1.00 | 1.00 | 43.0 |

The GCS pattern persists across LLM backends: linear mode fails under shift regardless of which model generates the summary (Llama3: 0.36, Mistral: 0.46), while graph mode maintains 1.00 for both. Mistral achieves slightly higher linear-mode GCS (0.46 vs 0.36), suggesting some models are marginally more likely to produce compliance language from degraded context, but the difference is not sufficient to replace architectural safety mechanisms. Mistral runs approximately 30% slower than Llama3 on this hardware. Figure 2 summarizes the GCS comparison across all tested configurations under institutional shift.

**Figure 2:**
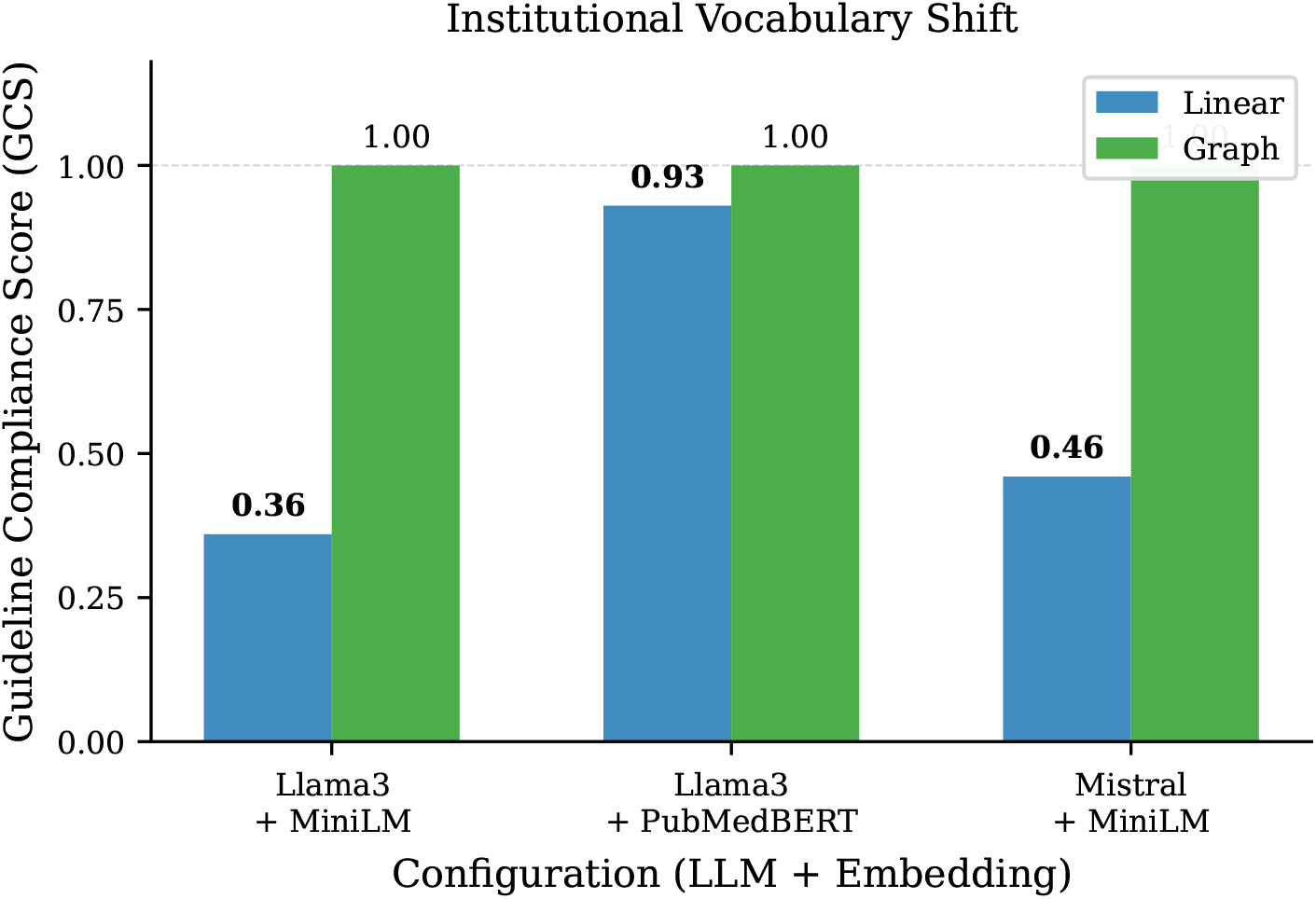
GCS under institutional vocabulary shift across three configurations. Linear mode fails in all cases (0.36–0.93 depending on embedding model), while graph mode maintains 1.00 regardless of embedding or LLM backend. PubMedBERT narrows the gap but does not close it.

Figure 3 shows the SFI-latency relationship across modes, with point size encoding compliance.

**Figure 3:**
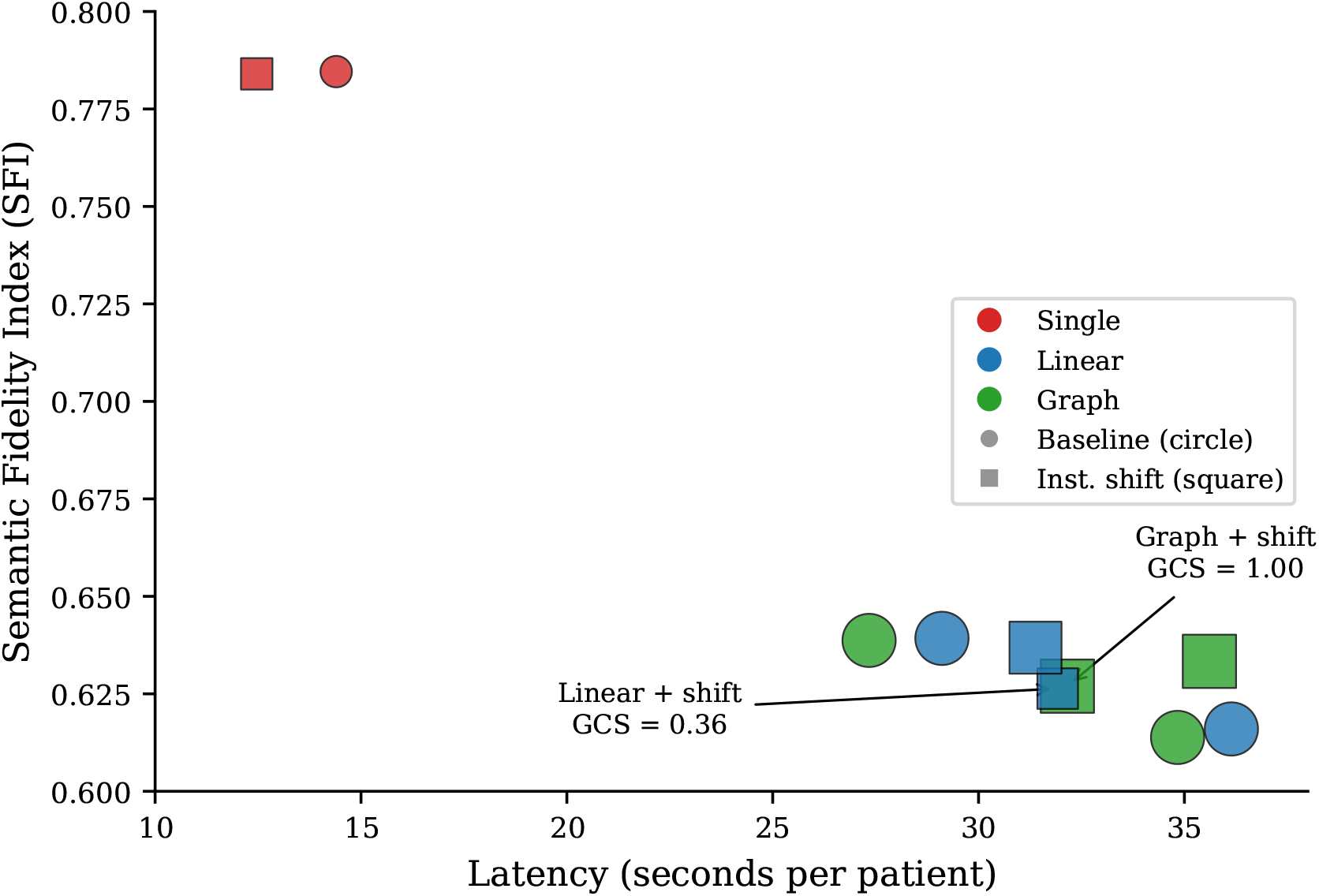
Semantic Fidelity Index versus latency per patient. Point size encodes GCS. Clusters of same-colored points represent multiple embedding model and LLM backend configurations for the same architecture mode, showing that the compliance pattern is stable across configurations. Under institutional shift, linear and graph modes occupy similar positions in SFI and latency but differ in compliance: the linear point is small (GCS = 0.36) while the graph point is large (GCS = 1.00).

## 5 Discussion

### Architecture determines compliance more than model quality

The central finding is that routing audit output back into the generation loop, rather than computing it post-hoc, is the single design decision that separates compliance from failure under shift. In linear mode, the auditor catches every contradiction (FCCR = 1.0). The problem is not detection. The problem is that the summary agent generates its output without ever seeing those findings. It writes a clinically plausible note, unaware that the guidelines it retrieved are the wrong ones. Graph mode closes that gap: the auditor’s output becomes input to the next iteration of the summary agent, so contradictions end up acknowledged in the final text.

This distinction matters for system designers. Adding an auditor to a pipeline is not enough. If the auditor’s output does not reach the component that produces patient-facing text, detection has no effect on compliance.

### Why PubMedBERT still misses 7% of patients

When we replaced MiniLM with PubMed-BERT, the linear pipeline’s compliance rose from 0.36 to 0.93. Better retrieval helps considerably. But the 7% residual failure has a specific cause. Our institutional vocabulary transformation includes substitutions like “eGFR” to “estimated renal clearance rate.” PubMedBERT handles most of these because it was trained on biomedical literature where synonymy is common. But compound substitutions, where two or three terms in a single sentence are replaced simultaneously (“metformin” becomes “biguanide class agent” in the same passage where “eGFR” becomes “estimated renal clearance rate”), push the cosine similarity below PubMedBERT’s retrieval threshold for approximately 1 in 14 patients.

In clinical terms: for every 500 T2DM patients with renal impairment passing through a RAG-based medication review, roughly 35 would receive a summary that fails to flag an active metformin contraindication. That is a non-trivial harm rate for a safety-critical application. The graph architecture eliminates it.

### Cross-model generalization

Mistral-7B and Llama3-8B share no training data, use different attention mechanisms (grouped-query vs. standard multi-head), and exhibit different instruction-following characteristics. Despite this, they produce the same GCS pattern: linear fails under shift, graph succeeds. Mistral’s linear GCS (0.46) is slightly higher than Llama3’s (0.36), which suggests Mistral is marginally more inclined to produce compliance-related phrasing even from degraded retrieval context. But 0.46 is still a failure rate of 54% on the affected subpopulation. No amount of model improvement within this size class closes the gap that architecture closes.

### Latency and clinical workflow fit

Graph mode costs 32 seconds per patient under institutional shift, compared to 12.5 seconds for single-agent and 31.9 seconds for linear. The safety loop adds only 3–5 seconds over linear mode when no contradictions are found (baseline conditions). Under shift, the loop iterates 2–3 times before converging or reaching *L*_*max*_, adding roughly 6 seconds.

Where does this fit in practice? Overnight batch reconciliation (processing a ward’s discharge summaries) tolerates minutes per patient. A pharmacist queue for formulary audit tolerates 30–60 seconds. Point-of-care decision support during a clinic visit does not tolerate 32 seconds. For that last case, either the safety floor would need pre-computation (running the audit during chart load, before the clinician requests a summary) or the loop depth would need dynamic adjustment based on time budget.

### Clinical implications for deployment

Hospitals considering RAG-based medication review systems face a concrete design choice. If they use a linear pipeline with domain-specific embeddings (the most common architecture in production), our data predicts that 7% of patients affected by institutional vocabulary differences will receive summaries with undetected contraindications. This is not a hypothetical: hospital mergers, EHR migrations, and formulary committee updates all produce exactly the kind of vocabulary shift we simulate.

The safety floor adds latency but eliminates this gap entirely, with no dependency on embedding model quality. For high-acuity populations (CKD stage 4, polypharmacy), the compliance guarantee may justify the extra seconds. For routine refill reviews, linear mode with PubMedBERT may be acceptable.

### Comparison to prior work

Prior evaluations of RAG failure modes [13, 26] have focused on retrieval degradation as an end in itself: does the system return the right documents? Xiong et al. found that retriever choice can swing downstream accuracy by up to 18 percentage points, confirming that retrieval quality matters. But our results show that retrieval failure is necessary but not sufficient for clinical harm. The clinical outcome depends on what happens downstream. A pipeline can retrieve the wrong document and still produce a safe output if its architecture routes contradictions back through a verification loop. Recent work on LLM hallucination in clinical summaries found rates of 1.47% per sentence across 12,999 clinician-annotated outputs [12]; our concern is different, because our failures are not random hallucinations but systematic omissions caused by shifted retrieval, which are harder to detect and more consistently harmful. Hegselmann et al. [32] showed that fine-tuning on hallucination-free data reduces errors from 2.6 to 1.55 per summary, but this addresses generation quality, not the architectural question of whether safety findings reach the generator.

Multi-agent reliability work [15, 18] has focused on coordination failures (agents disagreeing, infinite loops, or cascading hallucinations). We show a subtler failure mode: agents working correctly in isolation but the architecture failing to connect their outputs. The auditor in linear mode is correct. The summary agent is correct given its input. The system fails because information does not flow between them.

## 6 Limitations

1. **Synthetic data**. Patient profiles are programmatically generated. While distributions approximate real T2DM+CKD populations (Table 1), the data lacks production EHR noise, missingness, and heterogeneity. Phase 3 will validate on MIMIC-IV discharge notes under IRB approval.
2. **Single disease domain**. Scoped to T2DM with renal comorbidities. Generalizability to oncology, cardiology, or multi-system disease remains untested.
3. **Deterministic auditor**. Rule-based contradiction detection provides experimental control but underestimates production LLM auditor capabilities. Phase 2 will use an LLM-based auditor.
4. **Domain-specific embedding trade-off**. PubMedBERT reduces shift visibility, meaning our general-purpose model choice maximizes experimental discriminative power. Production systems should use biomedical embeddings combined with the safety floor.
5. **Model scale**. 7-8B parameter models on 36 GB unified memory. Larger models (70B+) may show different error patterns under shift.

## 7 Conclusion

Stateful multi-agent graphs with programmatic safety floors maintain full guideline compliance under institutional vocabulary shift that causes linear RAG pipelines to drop to 36% compliance. The key architectural decision is whether audit findings are routed back to the summary agent (graph mode) or computed post-hoc without influencing the output (linear mode). This pattern holds across embedding models and LLM backends, confirming it is an architectural property rather than a model-specific artifact. The framework runs on consumer hardware with no external API dependencies and is reproducible via a single command.

### Future work

(1) Phase 2 experiments with LLM-based parser/auditor. (2) MIMIC-IV validation under IRB. (3) Dynamic graph pruning for latency-critical workflows. (4) Multi-domain extension (cardiology, oncology).

## Data Availability

All data produced in the present work are contained in the manuscript. The source code and synthetic patient generation pipeline are available at https://github.com/Behordeun/clinicalshift

## Author contributions

M.A.S. conceived the study, designed and implemented the experimental pipeline, ran experiments, and drafted the manuscript. B.F.O. contributed to clinical domain modeling and manuscript revision.

H.S. contributed to manuscript revision.

## Data availability

All data are synthetically generated and reproducible via the published codebase. No real patient data were used.

## Ethics approval

Not required (synthetic data only, no human subjects).

## Footnotes

1 Code: https://github.com/Behordeun/clinicalshift

